# Tocilizumab, netakimab, and baricitinib in patients with mild-to-moderate COVID-19: a retrospective cohort study

**DOI:** 10.1101/2022.03.31.22269942

**Authors:** E.A. Bryushkova, V.D. Skatova, Z.Y. Mutovina, A.I. Zagrebneva, D.S. Fomina, T.S. Kruglova, A. Akopyan, I.D. Strazhesko, S. Lukyanov, O.N. Tkacheva, M.A. Lysenko, D.M. Chudakov

## Abstract

**Background:** The aim of the study was to assess inflammatory markers and clinical outcomes in adult patients admitted to hospital with mild-to-moderate COVID-19 and treated with targeted immunosuppressive therapy using anti-IL-17A (netakimab), anti-IL-6R (tocilizumab), or JAK1/JAK2 inhibitor (baricitinib) or with standard-of-care (SOC) therapy.

**Methods:** The retrospective, observational cohort study included 154 adults hospitalized between February and August, 2020 with RT-PCR-confirmed SARS-CoV-2 with National Early Warning Score2 (NEWS2) < 7 and C-reactive protein (CRP) levels ≤ 140 mg/L on the day of the start of the therapy or observation. Patients were divided into the following groups: I) 4 mg baricitinib, 1 or 2 times a day for an average of 5 days (n = 38); II) 120 mg netakimab, one dose (n = 48); III) 400 mg tocilizumab, one dose (n = 34), IV) SOC: hydroxychloroquine, antiviral, antibacterial, anticoagulant, and dexamethasone (n = 34).

**Findings:** CRP levels significantly decreased after 72 h in the tocilizumab (p = 1 × 10^−5^) and netakimab (p = 8 × 10^−4^) groups and remained low after 120 h. The effect was stronger with tocilizumab compared to other groups (p = 0.028). A significant decrease in lactate dehydrogenase (LDH) levels was observed 72 h after netakimab therapy (p = 0.029). NEWS2 scores significantly improved 72 h after tocilizumab (p = 6.8 × 10^−5^) and netakimab (p = 0.01) therapy, and 120 h after the start of tocilizumab (p = 8.6 × 10^−5^), netakimab (p = 0.001), or baricitinib (p = 4.6 × 10^−4^) therapy, but not in the SOC group. Blood neutrophil counts (p = 6.4 × 10^−4^) and neutrophil-to-lymphocyte ratios (p = 0.006) significantly increased 72 h after netakimab therapy and remained high after 120 h. The percentage of patients discharged 5-7 days after the start of therapy was higher in the tocilizumab (44.1%) and netakimab (41.7%) groups than in the baricitinib (31.6%) and SOC (23.5%) groups. Compared to SOC (3/34, 8.8%), mortality was lower in netakimab (0/48, 0%, RR=0.1 (95% CI: 0.0054 to 1.91)), tocilizumab (0/34, 0%, RR=0.14 (95% CI: 0.0077 to 2.67)), and baricitinib (1/38, 2.6%, RR=0.3 (95% CI: 0.033 to 2.73)) groups.

**Interpretation:** In hospitalized patients with mild-to-moderate COVID-19, anti-IL-17A or anti-IL-6R therapy were superior or comparable to the JAK1/JAK2 inhibitor, and all three were superior to SOC. Whereas previous studies did not demonstrate significant benefit of anti-IL-17A therapy for severe COVID-19, our data suggest that such therapy could be a rational choice for mild-to-moderate disease, considering the generally high safety profile of IL-17A blockers. The significant increase in blood neutrophil counts in the netakimab group may reflect efflux of neutrophils from inflamed tissues. We therefore hypothesize that neutrophil count and neutrophil-to-lymphocyte ratio could serve as markers of therapeutic efficiency for IL-17A-blocking antibodies in the context of active inflammation.

## Introduction

SARS-CoV-2 remains a pandemic pathogen with a high fatality rate, and efficient clinical protocols are still under development. After initial viral replication, the next and often most devastating stage of disease is driven by excessive or mistargeted host inflammatory response. Severely ill patients exhibit increased blood levels of interleukin (IL)-6, IL-1β, IL-2, IL-8, and IL-17, tumor necrosis factor α (TNF-α), granulocyte colony-stimulating factor (G-CSF), and interferon (IFN)-γ^1-5^. Increased IL-6 correlates with increased C-reactive protein (CRP) and both are elevated in non-survivors relative to survivors^6,7^. As such, the high potential of immunosuppressive therapies currently applied for rheumatologic disorders—ranging from dexamethasone and glucocorticosteroids to JAK inhibitors and selective cytokine inhibitors—as a treatment option for COVID-19 is now widely accepted^8-13^.

This study compares three therapies affecting linked inflammatory pathways.

Tocilizumab^14^ is an anti-IL-6R monoclonal antibody that blocks IL-6 signaling and trans-signaling^15^. This suppresses the pleiotropic effects of IL-6, which include activation of acute-phase protein production, promotion of Th17 and Th2 helper T cells, and suppression of Th1 helper T cell differentiation^16^. Tocilizumab has previously been used to treat severe rheumatological diseases and cytokine release syndrome in the context of chimeric antigen receptor (CAR) T cell therapy^17,18^. For this latter reason, many clinical centers have initially opted to apply tocilizumab to block cytokine storm induced by severe COVID-19^11,13,19,20^; currently, such treatment is recommended for hospitalized patients who are receiving systemic corticosteroids and require supplemental oxygen, non-invasive or invasive mechanical ventilation, or extracorporeal membrane oxygenation (https://www.fda.gov/news-events/press-announcements/coronavirus-covid-19-update-fda-authorizes-drug-treatment-covid-19).

Netakimab is an anti-IL-17A monoclonal antibody recommended for treatment of moderate-to-severe plaque psoriasis^21^, ankylosing spondylitis^22^ (ClinicalTrials.gov: NCT03447704) and psoriatic arthritis^23^ (ClinicalTrials.gov: NCT03598751). IL-17, which is produced by Th17, Tc17, and other clonal type 3-programmed RORγt-positive lymphoid cells^24^, triggers production of pro-inflammatory cytokines including granulopoiesis-inducing G-CSF, the systemic inflammatory cytokines IL-6, IL-1β, and TNFα, neutrophil-attracting IL-8, as well as tissue-disintegrating matrix metalloproteinases^25^. IL-6 in turn promotes the differentiation of Th17 cells, resulting in a positive feedback loop ^26^. There is substantial evidence that infection by coronaviruses, including MERS-CoV and SARS-CoV-2, directs the immune response along the Th17 pathway^27-31^, and IL-17 may represent one of the key mediators of tissue damage, acute respiratory distress syndrome (ARDS), and cytokine storm in such viral infections ^31-38^.

Baricitinib is a small molecule JAK1/JAK2 inhibitor used for the treatment of rheumatoid arthritis. Baricitinib affects intracellular signaling by decreasing the level of phosphorylated STAT proteins, thereby blocking the proinflammatory signals IL-6, IL-12, IL-23 and IFN-γ^39^. Baricitinib exerts a variety of immunosuppressive effects, including suppression of the differentiation of Th1 and Th17 cells^40^. This outcome links its mode of action with anti-IL-17A and anti-IL-6/IL-6R monoclonal antibodies. Based on positive clinical data^41-43^, baricitinib has been approved for COVID-19 treatment by the European Commission and FDA. The aim of this study was to assess inflammatory markers and clinical outcomes in hospitalized, mild-to-moderate COVID-19 patients treated with these three agents—netakimab, tocilizumab, and baricitinib—or with standard of care (SOC) therapy without targeted immunosuppression.

## Methods

### Study design and participants

This retrospective, observational cohort study was conducted in COVID-19 care units at two tertiary hospitals in Moscow (City Clinical Hospital No. 52 of the Moscow Healthcare Department, and the Russian Gerontology Research and Clinical Center at Pirogov Russian National Research Medical University) on patients with mild or moderate COVID-19 pneumonia. Both centers contributed data on baricitinib, netakimab, tocilizumab, and SOC treatment. The study participants were adults with RT-PCR-confirmed SARS-CoV-2 infection, who were admitted to the hospital between February and August 2020. Eligible patients had symptoms of acute respiratory infection (fever, muscle pain, cough) and radiologically-defined viral pneumonia as assessed by computed tomography.

Patients were excluded if their National Early Warning Score 2 (NEWS2) metric—which falls on a scale of 0 to 20 based on physiological variables including blood pressure, heart rate, respiratory rate, temperature, oxygen saturation, and level of consciousness^44-46^—was unknown or higher than 6 points at the start therapy day. Patients were also excluded if their serum CRP levels were higher than 140 mg/l, if there was no information about death or hospital discharge, if they were receiving combination or non-standard regimens of the investigated therapies, if therapy started later than 72 hours after admission to the hospital, or if they had severe haematological, renal, or liver function impairment or evidence of concomitant bacterial infections. All patients who received tocilizumab, netakimab, or baricitinib provided written informed consent. The study was approved by the local ethical committee of the City Clinical Hospital No.52.

### Procedures

All patients received SOC treatment at the time of hospital admission according to the corresponding COVID-19 Russian guidelines 2020 (Interim guidelines for the prevention, diagnosis and treatment of novel coronavirus infection (COVID-19). Version 4 (03/27/2020) https://static-0.minzdrav.gov.ru/system/attachments/attaches/000/049/881/original/COVID19_recomend_v4.pdf). In practice, however, SOC treatment varied for different patients, and situationally included antiviral, antibacterial, anticoagulant, dexamethasone treatment or convalescent plasma transfusion according to indications and contraindications. SOC treatment usually included hydroxychloroquine (400 mg twice on day 1, followed by 200 mg twice per day on days 5–10), azithromycin (500 mg once per day for 5 days), lopinavir–ritonavir (400/100 mg twice per day for 14 days), and low molecular-weight heparin according to indicators. In addition to SOC treatment, patients also received tocilizumab or baricitinib or netakimab treatment. Netakimab was administered by a single subcutaneous injection of 120 mg, tocilizumab was administered by a single intravenous dose of 400 mg, and baricitinib was administered at a dose of 4 mg once or twice per day for 3–8 days (5 days on average).

### Outcomes

The primary outcomes of the study were the biochemical inflammatory markers CRP (mg/L) and lactic dehydrogenase (LDH, U/L), absolute neutrophil count (ANC) and absolute lymphocyte count (ALC) in blood (× 10^9^/L), and the neutrophil-to-lymphocyte ratio (NLR). Secondary outcomes were the NEWS2 score, time until discharge from the hospital, and mortality rate. All parameters were estimated at baseline, 72 h after initiation of treatment, and 120 h after initiation of treatment.

### Statistical analysis

Continuous variables were expressed as median (IQR) and compared by Kruskal Wallis test. At baseline (0 h), we assessed the similarity of NEWS2, LDH, and CRP distribution in the four cohorts using the Kruskal-Wallis and Dunn test (**Table 2**). For paired comparisons between two time points, we used the Wilcoxon test. P values ≤ 0.05 were considered significant, all P values are shown with adjustment for multiple testing using Bonferroni correction. Analyses were done by R version 1.2.5. In order to extract maximal statistical information, we independently assessed the individual dynamics of parameters between baseline and 72 h (Δ0-72) and between baseline and 120 h (Δ0-120), including all patients for which the parameter of interest was known at both analyzed time points (**Fig. 1, Supplementary Table S1**). As such, the number of patients included in each analysis differs. Additionally, the individual dynamics of CRP, LDH, NEWS2 score, and blood cell counts was also assessed for patients for whom the results of sequential measurements were available for all three timepoints (**Fig. 2, Supplementary Fig. S3, Supplementary Table S2**). Again, the number of patients included in each analysis differed.

**Figure 1.**
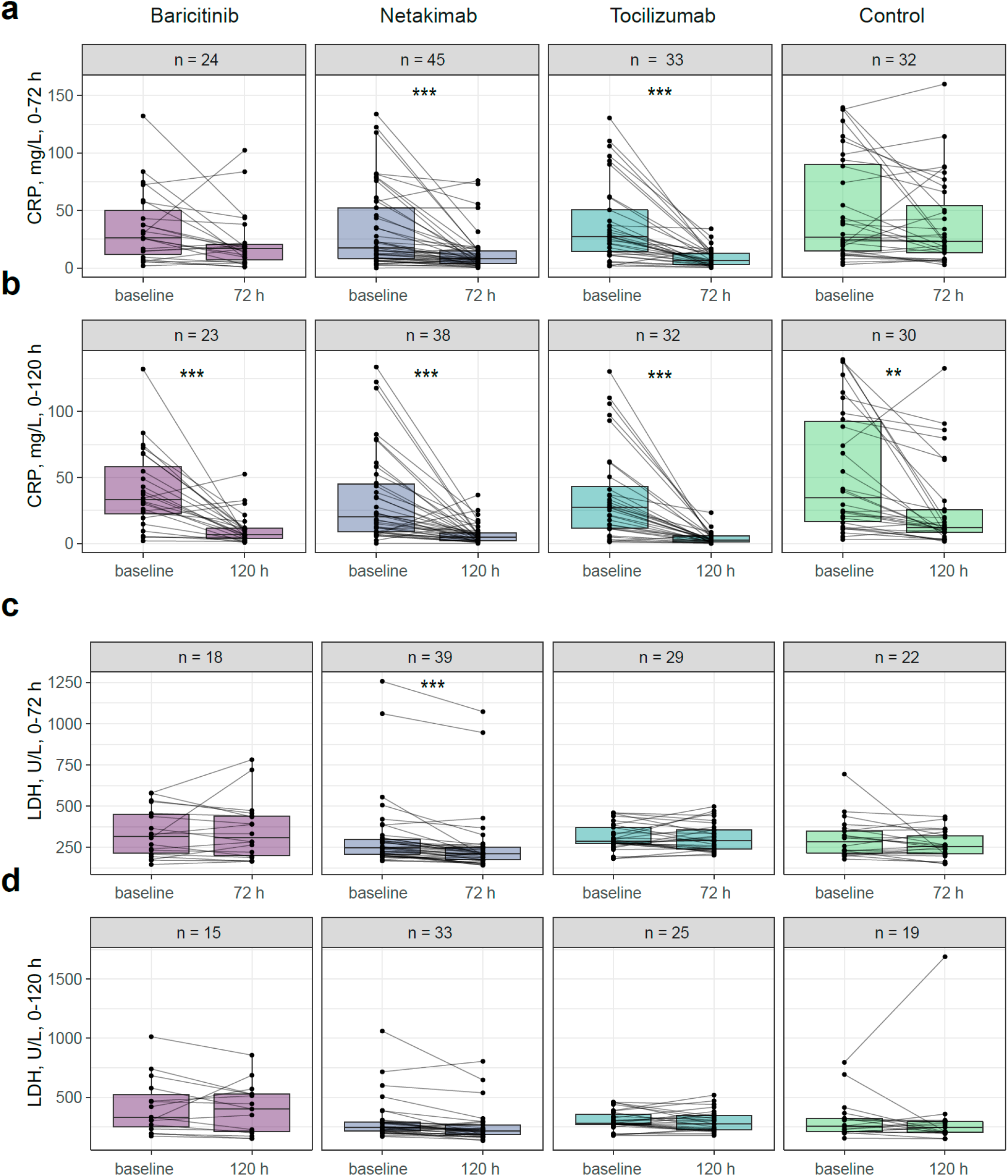
Changes in CRP and LDH levels over time. Measured levels of C-reactive protein (CRP) at baseline versus **a**, 72 h or **b**, 120 h. Measured levels of lactate dehydrogenase (LDH) at baseline versus **c**, 72 h or **d**,120 h. Dots show measurements for individual patients. Wilcoxon test: ∗∗p < 0.01, ∗∗∗p < 0.001. n = number of patients with available values of the corresponding parameters.

**Figure 2.**
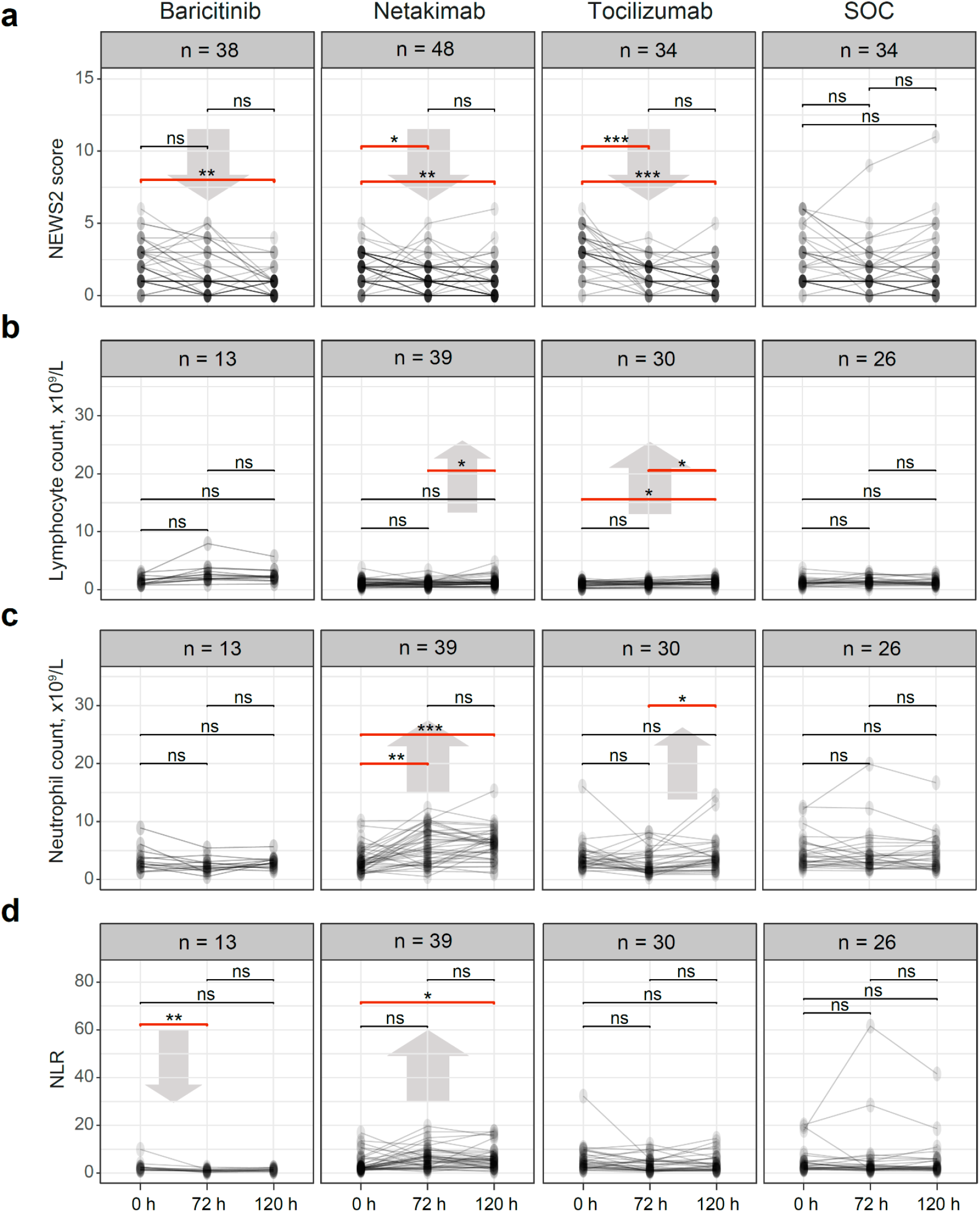
Individual dynamics of blood parameters. **a**. NEWS2 scores. **b**. Absolute lymphocyte counts. **c**. Absolute neutrophil counts (ANC). **d**. Neutrophil-to-lymphocyte ratio (NLR). Wilcoxon test: ∗p < 0,05, ∗∗p < 0.01, ∗∗∗p < 0.001. ns = non-significant. n = number of patients with available values of the corresponding parameters.

## Results

### Baseline patient characteristics

A total of 154 patients with mild-to-moderate COVID19 were included in the study to receive baricitinib (n = 38), netakimab (n = 48), tocilizumab (n = 34), or SOC (n = 34). Baseline characteristics of patients in the compared groups are summarized in **Tables 1** and **2**. Although the numbers of mild and moderate patients were roughly comparable in all groups, the Kruskal-Wallis and Dunn tests showed a significant difference in initial NEWS2 scores for tocilizumab compared to the netakimab group, while there were no significant differences between the baricitinib, netakimab, and SOC groups. Differences in LDH levels and NLR between groups were also significant at the start of the therapy (p = 0.04 and 0.012, respectively; **Table 2, Supplementary Fig. 1a**).

**Table 1.** Baseline patient characteristics.

| Parameter |  | Baricitinib<br>(n=38) | Netakimab<br>(n=48) | Tocilizumab<br>(n=34) | SOC<br>(n=34) |
| --- | --- | --- | --- | --- | --- |
| age, years, mean [SD] |  | 60.7 [12.8] | 63.4 [12.7] | 60.2 [8.7] | 66.1 [13.4] |
| sex, % (N) | male | 42 (16) | 48 (23) | 47 (16) | 32 (11) |
|  | female | 58 (22) | 52 (25) | 53 (18) | 68 (23) |
| days in hospital<br>before start of<br>therapy,% (N) | 0 | 34 (13) | 58 (28) | 82 (28) | - |
|  | 1 | 55 (21) | 38 (18) | 18 (6) |  |
|  | 2 | 11 (4) | 4 (2) | 0 |  |
| NEWS-2, %<br>(N) | 0-4 | 82 (31) | 92 (44) | 85 (29) | 79 (27) |
|  | 5-6 | 18 (7) | 8 (5) | 15 (5) | 21 (7) |
| Hydroxychloroquine treatment,<br>% (N) | yes | 68 (26) | 10 (5) | 35 (12) | 91 (31) |
|  | no | 32 (12) | 90 (43) | 65 (22) | 9 (3) |

Data are mean (SD) or n/N (%) where N = number; % = percentage; NEWS-2 = National Early Warning Score 2.

**Table 2.** The baseline clinical characteristics.

| Parameter | Baricitinib<br>(n = 38) | Netakimab<br>(n = 48) | Tocilizumab<br>(n = 34) | SOC<br>(n = 34) | p-value |
| --- | --- | --- | --- | --- | --- |
| **NEWS-2,**<br>**median (IQR)** | 2 (1–3) | 2 (1–3) | 3 (2–4) | 2 (1–3.75) | 0.007 |
| **ANC,**<br>**median (IQR)** | 5.05 (2.8–7.06) | 3,05 (1.92–4.95) | 3,25 (2.22–4.47) | 4.2 (2.85–6.47) | >0.05 |
| **ALC,**<br>**median (IQR)** | 1.65 (1.08–2.17) | 1 (0.72–1,6) | 1 (0.62–1.37) | 1.2 (0.87–1.52) | >0.05 |
| **NLR,**<br>**median (IQR)** | 2.42 (1.65–4,11) | 2.32 (1.77–5.46) | 3.94 (2.36–6.53) | 3.3 (1.95–5.65) | 0.011 |
| **CRP,**<br>**median (IQR)** | 28 (13.28–57.65) | 17.34 (8.32–52.36) | 27.39 (14.09–50.44) | 27.05 (14.98–89.91) | >0.05 |
| **LDH,**<br>**median (IQR)** | 327 (220.4–491.5) | 247.6 (209.4–298.8) | 286 (272.6–371.1) | 282.2 (216.4–350.4) | 0.039 |

NEWS-2 = National Early Warning Score 2; ANC = absolute neutrophil count ( $\times 10^9/L$ ); ALC = absolute lymphocyte count ( $\times 10^9/L$ ); NLR = neutrophil-to-lymphocyte ratio; CRP = C-reactive protein (mg/ml); LDH = lactic dehydrogenase (U/L); IQR = interquartile range. Evenness of parameters distribution between groups was examined with Kruskal-Wallis test. Paired comparisons between groups are shown in **Supplementary Fig. S1**.

### CRP and LDH

We independently assessed the individual dynamics of CRP and LDH for two separate datasets: patients with known parameters at baseline and 72 h (Δ0-72) and baseline and 120 h (Δ0-120) (**Supplementary Table 1**). This allowed us avoid excluding patients from the analysis who lacked CRP or LDH data at one of the time-points. CRP levels significantly decreased after 72 h in the tocilizumab (p = 1 × 10^−5^) and netakimab (p = 8 × 10^−4^) groups, but not in the baricitinib (p > 0.05) and SOC (p > 0.05) groups (**Fig. 1a**). The amplitude of this effect was higher in the tocilizumab group compared to the other groups (p = 0.028, **Supplementary Fig. S2**). After 120 h, CRP levels significantly decreased in all groups, but remained high for some individual patients in the SOC group (**Fig. 1b**). We only observed significant reduction of LDH levels in the netakimab group after 72 h (p = 0.029, **Fig. 1c,d**). The extent of LDH reduction did not significantly differ between groups after either 72 or 120 h (p > 0.05, **Supplementary Fig. S2c, d**).

We also assessed the individual dynamics of CRP and LDH for those patients for whom sequential measurements were available for all three time-points. For the vast majority of patients in the netakimab and tocilizumab groups, CRP decreased over time, while more heterogeneous dynamics were observed in the baricitinib and SOC groups (**Supplementary Fig. S3**).

### NEWS2 and blood cell counts

NEWS2 scores and blood cell counts were available from all time-points for most patients (**Fig. 2, Supplementary Table 2**). In this analysis, NEWS2 scores significantly improved 72 h after tocilizumab and netakimab therapy, and 120 h after the start of tocilizumab, netakimab, or baricitinib therapy, but not in the SOC group. Absolute neutrophil counts (ANC) increased significantly and prominently 72 h after netakimab therapy and remained high after 120 h. Blood neutrophil-to-lymphocyte ratios (NLR) significantly decreased after 72 h in the baricitinib group, but increased 120 h after netakimab therapy. We also performed separate analysis of NEWS2 scores and blood cell counts for patients with known parameters at baseline and 72 h (Δ0-72) and baseline and 120 h (Δ0-120), which allowed us to include more patients (**Supplementary Table 1**). This analysis confirmed the increase of ANC and NLR in the netakimab group at both 72 h and 120 h, as well as the decrease of NLR in the baricitinib group.

### Day of discharge and mortality data

Most patients were discharged from the hospital by the 14th day after initiation of therapy (**Table 3**), without significant differences between the groups (p > 0.05). The percentage of patients who were discharged 5–7 days after the start of therapy was higher in the tocilizumab (44.1% of patients) and netakimab (41.7%) groups relative to the baricitinib (31.6%) and SOC (23.5%) groups. Mortality was higher in the SOC (9%) and baricitinib (3%) groups than in tocilizumab (0%) and netakimab (0%) groups (**Table 3**).

**Table 3.** The outcome timepoints and the mortality level of patients.

| Group |  | Baricitinib<br>(n=38) | Netakimab<br>(n=48) | Tocilizumab<br>(n=34) | SOC<br>(n=34) | p-value |
| --- | --- | --- | --- | --- | --- | --- |
| Outcome,<br>% (N) | hospital<br>discharge | 97 (37) | 100 (48) | 100 (34) | 91 (31) | p >0.05* |
|  | death | 3 (1) | 0 (0) | 0 (0) | 9 (3) | - |
| Days in<br>hospital after<br>starting<br>therapy, %<br>(N) | ≤4 | 2.6 (1) | 4.2 (2) | 0 (0) | 0 (0) |  |
|  | 5–7 | 28.9 (11) | 37.5 (18) | 44.1 (15) | 23.5 (8) |  |
|  | 8–14 | 57.9 (22) | 47.9 (23) | 47.1 (16) | 70.6 (24) |  |
|  | ≥15 | 10.5 (4) | 10.4 (5) | 8.8 (3) | 5.9 (2) |  |

Kruskal-Wallis and Dunn test.

## Discussion

*Tocilizumab*. Tocilizumab was recommended on 6 July 2021 by the WHO for treatment of severe COVID-19. It was the second drug found to be effective against COVID-19 after dexamethasone, in September 2020 (https://www.who.int/news/item/06-07-2021-who-recommends-life-saving-interleukin-6-receptor-blockers-for-covid-19-and-urges-producers-to-join-efforts-to-rapidly-increase-access). Some previous data have suggested that tocilizumab might reduce the risk of invasive mechanical ventilation or death in patients with severe ^19^ or moderate-to-severe ^47^ COVID-19 pneumonia. However, these results were controversial, because other clinical trials found that tocilizumab was not effective for preventing intubation or death in moderately ill patients ^48,49^ (ClinicalTrials.gov, <u>NCT04356937</u>). The results of the COVACTA phase 3 clinical trial (ClinicalTrials.gov, <u>NCT04320615</u>) were published in summer 2020. This randomized controlled trial was conducted at 62 hospitals in nine countries (Canada, Denmark, France, Germany, Italy, the Netherlands, Spain, the United Kingdom, and the United States) and included 438 patients. All patients were hospitalized with COVID-19 pneumonia and received an intravenous infusion of 8 mg/kg tocilizumab (800 mg maximum, n = 294) or placebo (n = 144). The main outcomes were clinical status at days 14 and 28, mortality at day 28, and number of ventilator-free days by day 28. Tocilizumab treatment did not yield significantly better clinical status or lower mortality than placebo at 28 days. Adverse events were similar and well balanced in the two trial groups, although incidence of infection was lower in the tocilizumab group ^50^.

The 11-month RECOVERY trial (ClinicalTrials.gov, NCT04381936) included 4,116 patients who had hypoxia and evidence of inflammation (CRP ≥ 75 mg/L); 2,022 patients received 400–800 mg tocilizumab plus SOC therapy, with 2,094 patients only receiving SOC therapy. Almost all patients (82%) were also taking systemic steroids (*e*.*g*. dexamethasone). 28-day mortality was significantly lower in the tocilizumab group compared to the control group (31% vs 35%), along with a greater probability of discharge from hospital within 28 days (57% vs 50%) and lower risk of invasive mechanical ventilation or death (35% vs 42%)^13^. These results led to an update in WHO clinical guidelines and the inclusion of tocilizumab as a COVID-19 treatment.

Although the RECOVERY trial did not observe significant differences in the frequency of new cardiac arrhythmias between tocilizumab and control groups, and described only three reports of serious adverse reactions believed to be related to tocilizumab (otitis externa, *Staphylococcus aureus* bacteraemia, and lung abscess), many authors have mentioned significantly higher levels of other adverse effects of tocilizumab, including serious hepatic, pancreatic, and pulmonary reactions^51^, cases of neutropenia, superinfections, reactivation of latent infections^52^, and late-onset infections among patients receiving tocilizumab^53^. Nevertheless, the current consensus is that tocilizumab may be effective in severe COVID-19 disease, especially in patients with infection-related inflammation and lung damage^48^. In our study, tocilizumab produced a profound and stable decrease of CRP, and significant improvement in NEWS2 scores 72 h after therapy. The tocilizumab group had a higher percentage of patients discharged 5-7 days after the therapy, with lower mortality compared to the baricitinib and SOC—but not netakimab—groups.

*Baricitinib*. At the start of this study (April 2020), there were no recommendations on the use of baricitinib for the treatment of COVID-19. The motivation for its use was to suppress pathological inflammatory response. Previous clinical trials on healthy volunteers have demonstrated that maximal inhibition of cytokine-induced STAT3 phosphorylation was observed 1–2 h after administration of baricitinib at single oral doses of 1, 5, or 10 mg, with levels returning to baseline by 16–24 h^54^. Previous clinical studies of baricitinib effectiveness and safety for patients with rheumatoid arthritis did not show any significant difference between 4 mg and 8 mg doses. Treatment-related adverse events were observed in 63% and 67% of patients receiving the 4 mg and 8 mg dosage, respectively; the incidence of serious cases (including infection) was 35% and 40%, respectively^55^. Based on these results, patients were maintained at a stable concentration of baricitinib, dosed at 4 mg one or two times a day for 3–8 days (five days on average). As a whole, studies of long-term (2–5.5 years) administration of baricitinib for rheumatoid arthritis have indicated good tolerability. The most common side effects in patients treated with 4 mg/day baricitinib include risk of pneumonia, herpes zoster, and infections of the urogenital tract, as well as deterioration of biochemical parameters, neutropenia, lymphopenia, anemia, thrombocytosis, and elevation of liver enzymes (ALT, AST), lipids, and creatine phosphokinase. Long-term use of baricitinib also increases the probability of developing deep venous thrombosis (DVT) and pulmonary embolism (PE) in patients belonging to corresponding risk groups. Baricitinib is therefore not recommended for patients with active tuberculosis or other infections, as well as patients with ALC < 0.5 × 10^9^/L, ANC < 1.0 × 10^9^/L or Hb < 8 g/dL, and should be used with caution in patients with risk factors such as old age, obesity, history of DVT / PE, or use of selective COX-2 inhibitors^39^. There is no definitive answer as to whether the above-described side effects manifest after short-term (7–14 days) use of baricitinib for COVID-19. Retrospective studies at the University of Pisa (37 patients, 4 mg/day for 14 days) and Albacete Hospital (46 patients, 2 or 4 mg/day for 3–11 days) demonstrated that the beneficial effect of baricitinib became apparent at the fifth day of use and persisted until the end of observation (P < 0.0001). During that same time, transaminitis was observed in seven (19%) patients within 72 hours after starting baricitinib, and resulted in discontinuation of the drug in four cases; for the other three, the biochemical parameters stabilized on their own. In addition, five (14%) patients from Pisa got treatment-related infections. In the Albacete cohort, transaminitis was observed in 12 (26%) patients, but therapy was not terminated and the health indicators returned to normal regardless of the drug intake. Other side effects—probably associated with baricitinib treatment—were observed in nine (20%) patients^56^.

More extensive results were obtained recently from a phase 3 double-blind, randomized placebo-controlled study (ClinicalTrials.gov, NCT04421027), which enrolled 1,525 patients over 18 years old from 12 countries in Asia, Europe, North America, and South America with laboratory-confirmed COVID-19. Baricitinib or placebo was administered orally at a dose of 4 mg/day (or 2 mg/day for patients with baseline estimated glomerular filtration rate of 30–60 (mL/min)/1.73 m^2^) for 14 days or discharge, if it happened before 14 days. The baricitinib group achieved absolute risk reduction of five percentage points in all-cause mortality at 28 days, and 4.9 percentage points in all-cause mortality at 60 days. At the same time, the frequency of therapy-associated adverse effects was the same in the baricitinib and placebo groups. At least one treatment-related adverse event was observed for 45% of patients in the baricitinib group vs 44% in the placebo group, with serious adverse events described for 15% and 18% of participants, respectively. The combination of baricitinib with SOC did not lead to an increased risk of infections for this cohort of patients^42^.

*Netakimab*. Netakimab is a humanized IgG1 monoclonal antibody against IL-17a that was registered in Russia in spring 2020 for the treatment of psoriasis^21^, ankylosing spondylitis^22^ (ClinicalTrials.gov: NCT03447704;), and psoriatic arthritis (ClinicalTrials.gov: NCT03598751). Clinical trials suggest that netakimab is well tolerated. In a phase 3 clinical study of moderate-to-severe plaque psoriasis, PLANETA (ClinicalTrials.gov: NCT03390101) patients received netakimab 120 mg once every two or four weeks over the course of 12 weeks^21^. Treatment-emergent adverse events were registered in 9.4%, 10.7%, and 6.8% of patients receiving two-week, four-week, or placebo treatment, respectively (P > 0.05). All treatment-related adverse events were typical for IL-17 inhibitors: neutropenia, lymphopenia, and upper respiratory tract infections. The potential efficacy of netakimab for treating cytokine release syndrome in COVID-19 has been examined in at least two retrospective studies: in comparison with patients who did not receive anti-cytokine therapy or were treated by tocilizumab^57^, and in comparison with SOC therapy including hydroxychloroquine, azithromycin, low-molecular-weight heparins, and corticosteroids^58^. Both studies indicated that netakimab is safe. The first study included COVID-19 patients with CRP > 60 mg/L, and the authors observed decreased CRP, fibrinogen, creatinine, and body temperature, and increased lymphocyte and platelet counts and alanine-transferase activity in all groups of patients 7–10 days after the start of therapy. The decrease in CRP levels was greater in the netakimab and tocilizumab groups than in the control group, and there was no difference between the netakimab and tocilizumab groups. Patients who received netakimab had a better survival rate among all three groups. In the second study, NEWS2 and CRP scores improved significantly on day three after the start of therapy for the netakimab group compared to the SOC group, but there was no difference in LDH levels and mortality^58^. Unlike in our study, patients with severe disease were included, with a median NEWS2 score of 7 for the whole cohort.

A study investigating hospitalized patients with severe COVID-19 treated with another IL-17 blocking antibody, secukinumab, showed that the therapy was well tolerated and was not associated with increase of adverse events or secondary infections compared to the SOC group. While no significant benefit was shown for the severe patients, a significant decrease of the number of thromboembolic events was demonstrated (https://www.medrxiv.org/content/10.1101/2021.07.21.21260963v1).

In our study of mild-to-moderate COVID19 patients, the netakimab group was the only one in which a significant decrease of both CRP and LDH parameters was observed 72 h after the start of therapy. This group was also characterized by a higher percentage of patients discharged 5-7 days after the therapy, as well as zero mortality.

It is remarkable that we observed a significant increase in peripheral neutrophil counts specifically in the netakimab group (**Fig. 2**), in the context of decreasing CRP (**Fig. 1a,b**), LDH (**Fig. 1c,d**), and NEWS2 (**Fig. 2**) and a positive clinical outcome (**Table 3**). IL-17 triggers production of a range of pro-inflammatory cytokines and chemokines, including neutrophil-attracting IL-8 (Ref. ^25^); in ARDS patients with a systemic inflammatory response, alveolar levels of IL-17 are increased, and this is associated with organ dysfunction and increased alveolar neutrophils and alveolar permeability^33,34^. Therefore, our interpretation of these results is that blockage of IL-17 activity could lead to the partial loss of neutrophil chemotaxis to the inflamed lung tissues, and their efflux from inflamed tissues results in the observed increase in peripheral blood neutrophil counts. As such, the ANC and NLR parameters could paradoxically be considered as markers of therapeutic efficicacy for IL-17-blocking antibodies in the context of active inflammation.

## Conclusion

We conclude that the use of any of the three targeted immunosuppression regimens under consideration—tocilizumab, netakimab, or baricitinib—results in an improvement in CRP levels and a positive trend in NEWS2 score 120 h after the start of the therapy in patients with mild to moderate coronavirus infection relative to SOC therapy. However, our results indicate that tocilizumab or netakimab might be more effective in alleviating inflammation within the first 72 hours of therapy. Potential adverse events should probably be the key criteria for selecting immunosuppressive therapy for such patients, and considering the generally high safety profile of anti-IL-17 blockers^59^, those may represent the best option of the three for this category of patients. A prospective randomized trial will be necessary to verify our results, but we hope that our current observations will be useful in guiding therapy in individual cases based on the patient’s condition and potential risk of adverse events for each drug.

## Data Availability

All data produced in the present work are contained in the manuscript

## Acknowledgments

Supported by grant of the Ministry of Science and Higher Education of the Russian Federation № 075-15-2020-807. We are grateful to Michael Eisenstein for his valuable help in editing the manuscript.

## Supplementary data

**Supplementary Fig. S1.**
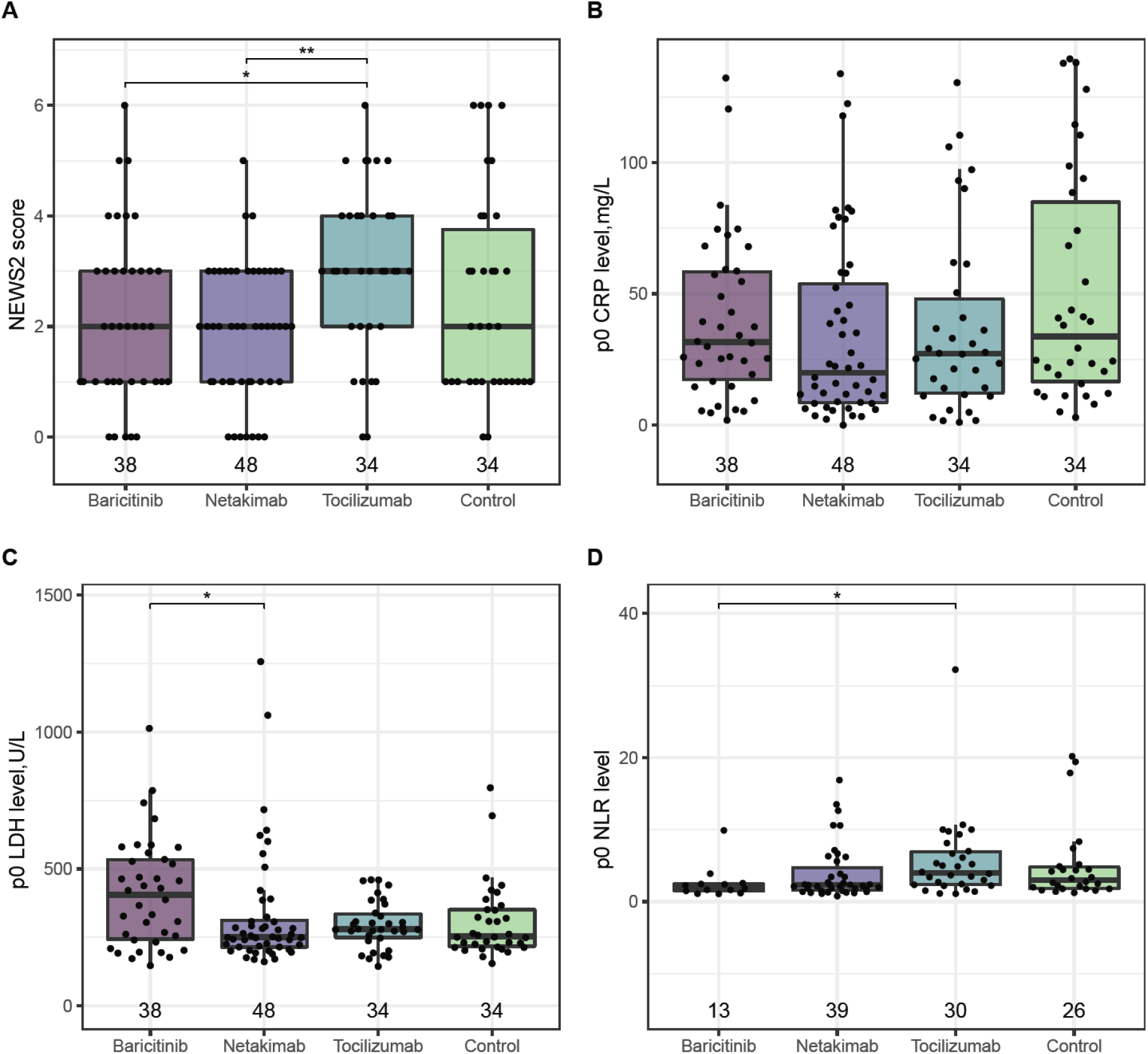
Distribution of the **a**, NEWS-2 score **b**, CRP level, **c**, LDH level, and **d**, NLR (D) at baseline. Kruskal-Wallis and Dunn test. *p <0.05, **p <0.01, ***p <0.001. Bonferroni adjustment method.

**Supplementary Fig. S2.**
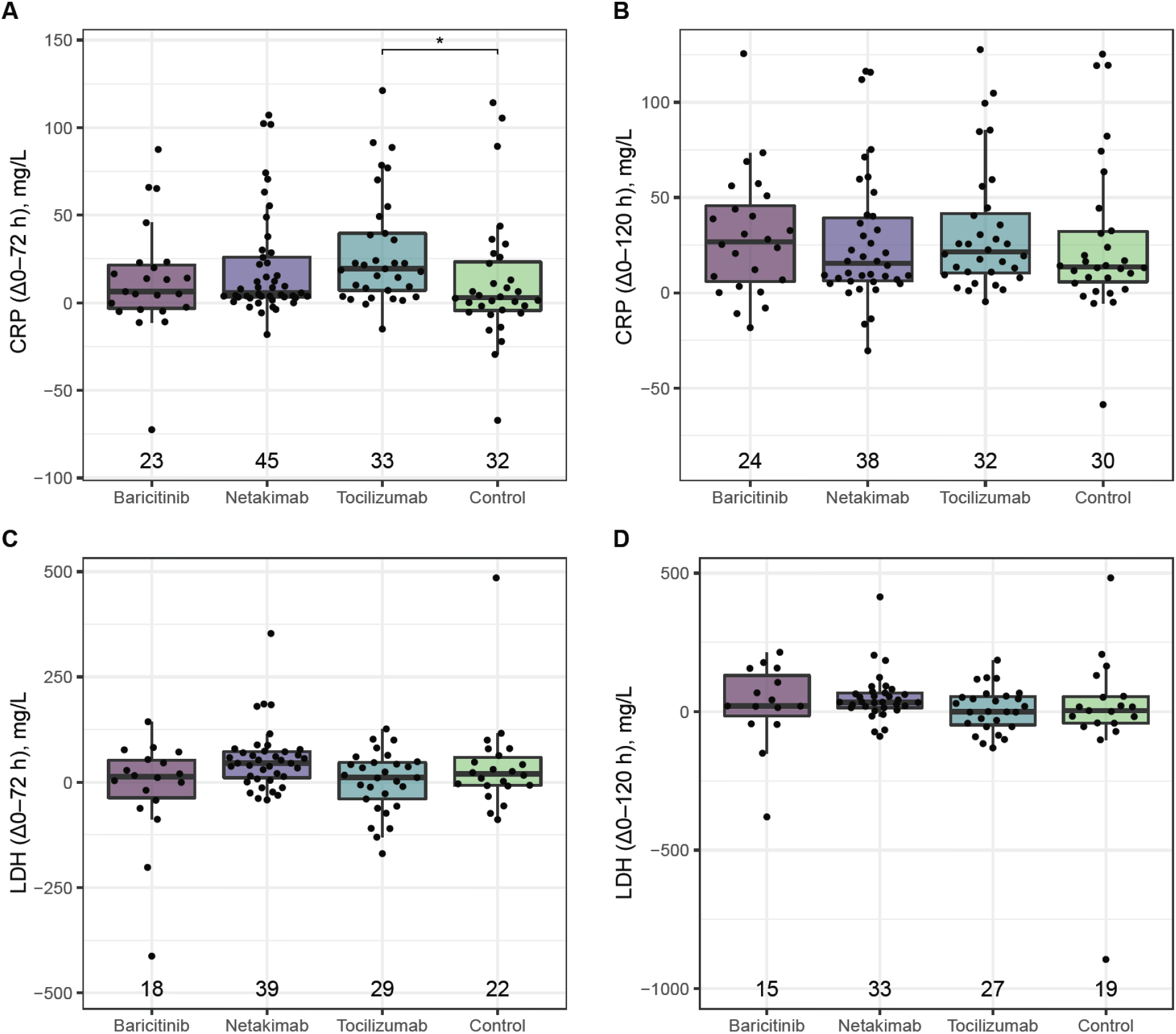
Changes in CRP and LDH levels 72 (A,C) or 120 hours (B,D) after the start of therapy. Kruskal-Wallis and Dunn test. *p <0.05. Number of patients with available data are shown along the x-axis.

**Supplementary Fig. S3.**
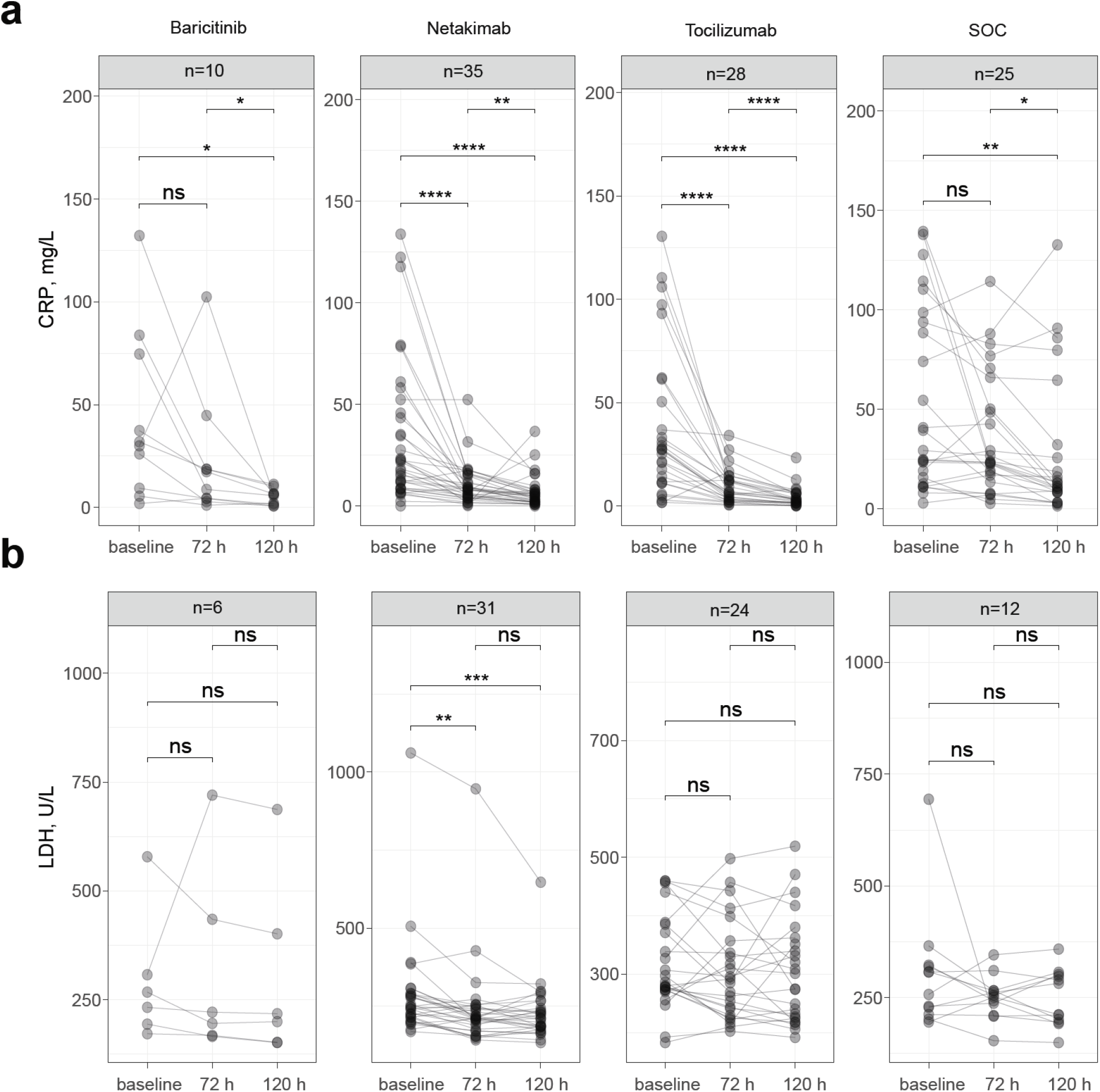
Individual dynamics of **a**, CRP and **b**, LDH across 120 h of treatment. Wilcoxon test. ∗p <0,05, ∗∗p <0,01, ∗∗∗p <0,001, ∗∗∗∗p <0,0001, ns = non-significant. n = number of patients with available data for all 3 time-points.

**Supplementary Table S1.**
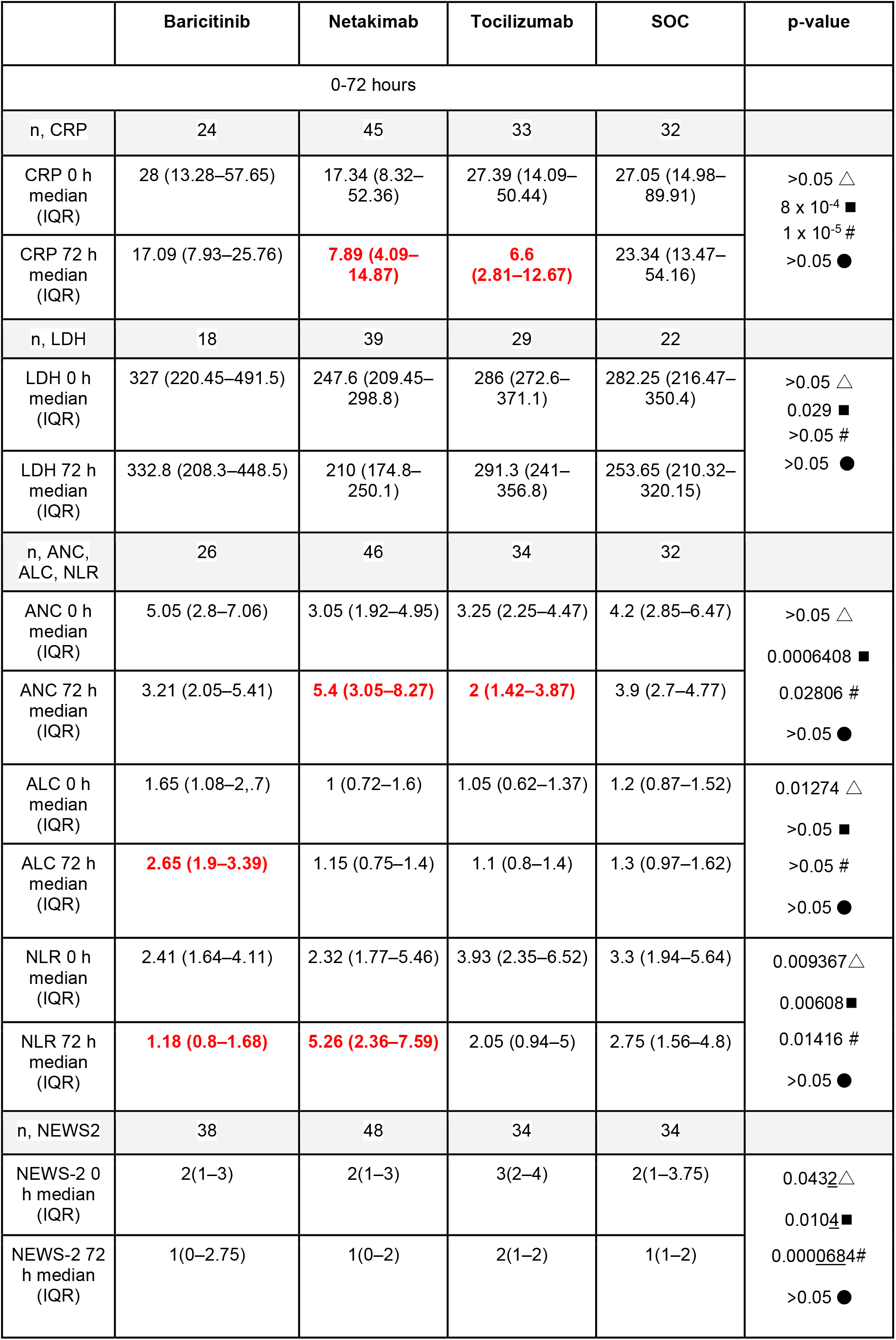
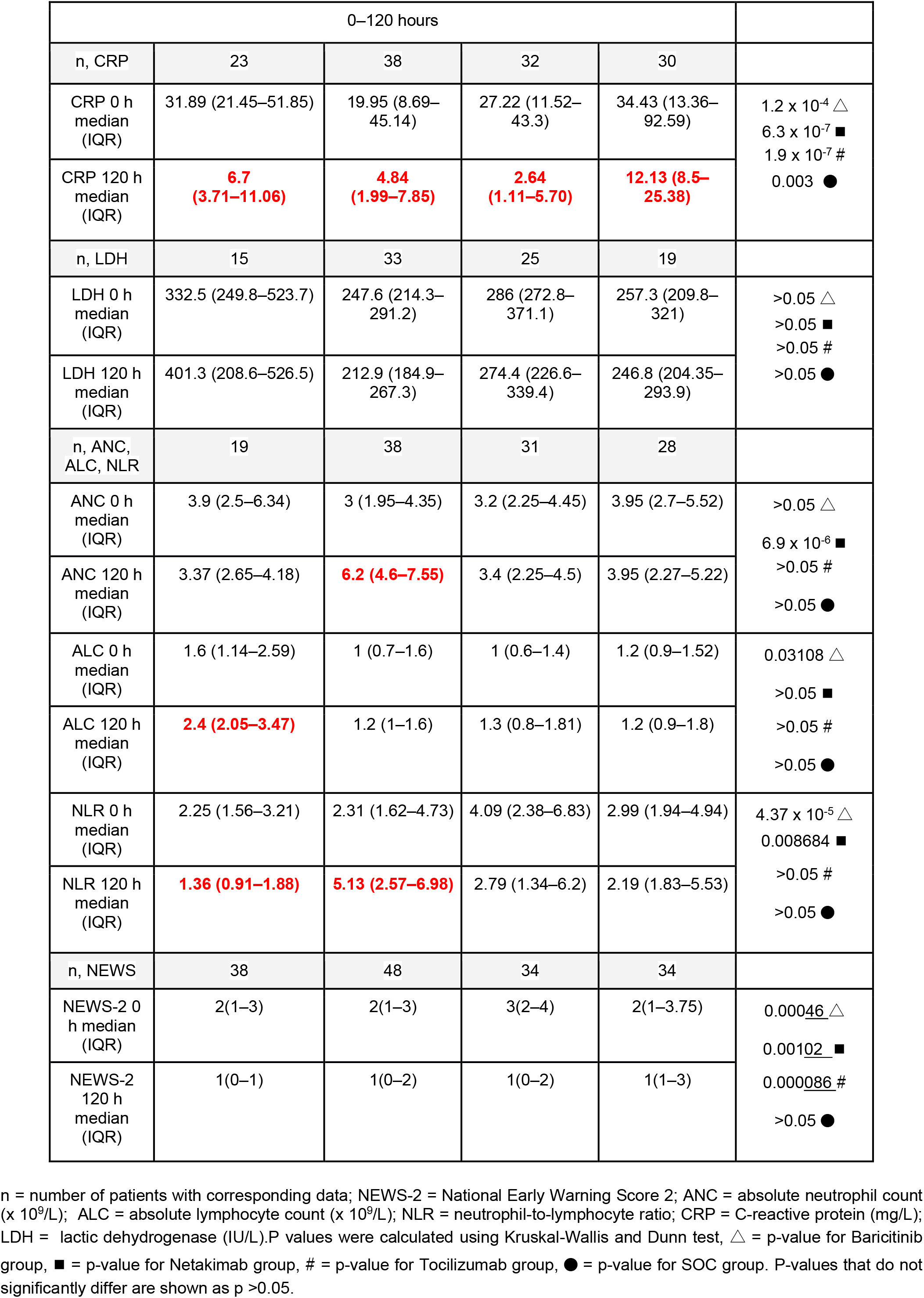
Clinical characteristics of patients from the Δ0–72 h and Δ0–120 h groups.

**Supplementary Table S2.** Clinical characteristics of patients at different timepoints.

|  | Baricitinib | Netakimab | Tocilizumab | SOC |
| --- | --- | --- | --- | --- |
| n, CRP | 10 | 35 | 28 | 25 |
| CRP 0 h<br>median (IQR) | 31.89 (14.6–72.4) | 17.76 (8.59–44.01) | 27.39 (12.87–45.68) | 27 (14.98–95.13) |
| CRP 72 h<br>median (IQR) | 18,46 (4.34–37.93) | **7.9 (3.98–12.55)** | **6.48 (2.52–12.4)** | 23.6 (16.09–67.23) |
| CRP 120 h<br>median (IQR) | 5.73 (2.1–6.7) | **4.72 (1.96–6.95)** | **2.7 (0.98–5.75)** | 12.13 (8.44–27.34) |
| n, LDH | 6 | 31 | 24 | 12 |
| LDH 0 h<br>median (IQR) | 267.3 (213.05–319.9) | 245.9 (214.25–286.8) | 280 (272.6–371.1) | 257.3 (212.5–319.2) |
| LDH 72 h<br>median (IQR) | 221.2 (181.65–383.75) | 211.6(182.7–244.9) | 285 (238.5–336) | 251.6 (120.1–265.7) |
| LDH 120 h<br>median (IQR) | 217.7(175.7–314.15) | 210.1 (183.95–240.25) | 274,4 (226.6–351.2) | 248,6 (201.6–298) |
| n, ANC, ALC, NLR | 13 | 39 | 30 | 26 |
| ANC 0 h<br>median (IQR) | 2.8 (2.2–3.9) | 3 (1.95–4.35) | 3.4 (2.22–4.3) | 3.95 (2.7–5.97) |
| ANC 72 h<br>median (IQR) | 2.2 (1.8–2.9) | **5.3 (2.9–8.4)** | 2.1 (1.35–4.12) | 3.65 (2.62–4.82) |
| ANC 120 h<br>median (IQR) | 2.9 (2.2–3.37) | **6.2 (4.6–7.55)** | 3.45 (2.47–4.7) | 3.65 (2.22–5.67) |
| ALC 0 h<br>median (IQR) | 1.5 (0.92–1.9) | 1 (0.7-1.5) | 0.95 (0.6–1.4) | 1.2 (0.9–1.57) |
| ALC 72 h<br>median (IQR) | **2.3 (1.9–3.2)** | 1.2 (0.75–1.4) | 1.05 (0.72–1.3) | 1.35 (1–1.9) |
| ALC 120 h<br>median (IQR) | **2.2 (2–2.4)** | 1.2 (1–1.6) | 1.3 (0.8–1.8) | 1.15 (0.9–1.75) |
| NLR 0 h<br>median (IQR) | 2.4 (1.48–3.87) | 2.32 (1.58–3.75) | 3.99 (2.35–6.91) | 2.56 (1.77–6.29) |
| NLR 72 h<br>median (IQR) | 1.31(0.96–2.44) | **4.85 (2.44–8.12)** | 2.81(1.04–5.04) | 2.75 (1.63–6.75) |
| NLR 120 h<br>median (IQR) | 1.57 (1.16–2.07) | **5.2 (2.26–7.07)** | 2.89 (1.38–6.3) | 3.41 (1.92–5.9) |

n = number of patients with data available; NEWS-2 = National Early Warning Score 2; ANC = absolute neutrophil count ( $\times 10^9/L$ ); ALC = absolute lymphocyte count ( $\times 10^9/L$ ); NLR = neutrophil-to-lymphocyte ratio; CRP = C-reactive protein (mg/L); LDH = lactic dehydrogenase (IU/L).

